# Risk factors for long COVID among healthcare workers, Brazil, 2020–2022

**DOI:** 10.1101/2023.01.03.22284043

**Authors:** Alexandre R. Marra, Vanderson Souza Sampaio, Mina Cintho Ozahata, Rafael Lopes Paixão da Silva, Anderson Brito, Marcelo Bragatte, Jorge Kalil, João Luiz Miraglia, Daniel Tavares Malheiros, Yang Guozhang, Vanessa Damazio Teich, Elivane da Silva Victor, João Renato Rebello Pinho, Adriana Cypriano, Laura Wanderly Vieira, Miria Polonio, Solange Miranda de Oliveira, Victória Catharina Volpe Ricardo, Aline Miho Maezato, Gustavo Yano Callado, Guilherme de Paula Pinto Schettino, Ketti Gleyzer de Oliveira, Rúbia Anita Ferraz Santana, Fernanda de Mello Malta, Deyvid Amgarten, Ana Laura Boechat, Takaaki Kobayashi, Eli Perencevich, Michael B. Edmond, Luiz Vicente Rizzo

**Author notes:** **<u>Corresponding author</u>**: Alexandre R. Marra, MD, University of Iowa, Department of Internal Medicine, 200 Hawkins Dr., C51-GH, Iowa City, IA 52242. **<u>Alternate corresponding author</u>**: Prof. Luiz Vicente Rizzo, MD, Hospital Israelita Albert Einstein, Instituto Israelita de Ensino e Pesquisa Albert Einstein, São Paulo, Brazil; Av. Albert Einstein, 627, Morumbi, São Paulo, Brazil - 05651-901.

## Abstract

**Objectives:** We aimed to determine risk factors for the development of long coronavirus disease (COVID) in healthcare workers (HCWs).

**Methods:** We conducted a case-control study among HCWs who had confirmed COVID-19 infection working in a Brazilian healthcare system between March 1, 2020 and July 15, 2022. Cases were defined as those having long COVID per the Centers for Disease Control and Prevention definition. Controls were defined as HCWs who had documented COVID-19 infection but did not develop long COVID. Multiple logistic regression was used to assess the association between exposure variables and long COVID during 180 days of follow-up.

**Results:** Of 7,051 HCWs diagnosed with COVID-19 infection, 1,933 (27.4%) who developed long COVID were compared to 5,118 (72.6%) who did not. The majority of those with long COVID (51.8%) had 3 or more symptoms. Factors associated with development of long COVID were female sex (OR 1.21 [CI95 1.05-1.39]), age (OR 1.01 [CI95 1.00-1.02]), and two or more COVID-19 infections (1.27 [CI95 1.07-1.50]). Those infected with the Delta variant (OR 0.30 [CI95 0.17-0.50]) or the Omicron variant (OR 0.49 [CI95 0.30-0.78]), and those receiving four COVID-19 vaccine doses prior to infection (OR 0.05 [CI95 0.01-0.19]) were significantly less likely to develop long COVID.

**Conclusions:** Long COVID can be prevalent among HCWs. We found that acquiring more than one COVID-19 infection was a major risk factor for long COVID, while maintenance of immunity via vaccination was highly protective.

## BACKGROUND

The majority of people infected by the SARS-CoV-2 virus tend to have complete resolution of their symptoms within a few days to a few weeks after infection, but some develop prolonged symptoms following acute infection [1, 2]. The post-coronavirus disease (COVID) conditions, also known as long COVID, according to the Centers for Disease Control and Prevention (CDC), include a wide range of new, returning, or ongoing health problems that people experience four or more weeks after first being infected with the virus that causes COVID-19 [1]. It has been estimated that approximately 200 million individuals have experienced these prolonged symptoms, with a global prevalence of 43% of the infected [2].

In the third year of the COVID-19 pandemic, 68.3% of the world population has received at least one dose of a COVID-19 vaccine. It has already been proven that in fully vaccinated populations, the vaccine effectiveness (VE) was high against infection (89.1%) and severe outcomes such as hospitalization (97.2%), admission to an intensive care unit (97.4%), and death (99.0%) [3]. However, despite the high effectiveness of the available vaccines [4-6], parts of the population remain susceptible to infection and long COVID [7-9]. Moreover, long COVID has been linked to more than 3,500 deaths emphasizing the need for prevention of infection [9].

Studies have shown that long COVID can follow both mild acute disease and the most severe forms [10-12]. Risk factors for long COVID in non-hospitalized adults include female sex, socioeconomic deprivation, obesity, and a wide range of comorbidities [12]. Healthcare workers (HCWs) have been identified as a more vulnerable group to infection, due to their high frequency of occupational exposure [13, 14]. Recent studies have shown that most vaccinated individuals report improvement of long COVID symptoms [15]. Among the few studies available, a cohort study that followed HCWs found that the number of vaccine doses was associated with a lower prevalence of long COVID [16]. Additional studies are needed to evaluate the effectiveness of COVID-19 vaccines against long COVID among HCWs [17-19]. In the present study, we aimed at analyzing epidemiological data and risk factors for the development of long COVID among HCWs in Brazil.

## METHODS

### Population and setting

This was a case-control study that included HCWs (aged ≥ 18 years) at Hospital Israelita Albert Einstein (HIAE) from March 1, 2020 to July 15, 2022. The HIAE is a Brazilian nonprofit healthcare, educational, and research organization, headquartered in the city of São Paulo, hat manages a diverse healthcare system ranging from primary healthcare to tertiary care services in the public and private healthcare sectors. It operates 40 healthcare units, mainly in the state of São Paulo, and in 2021 it had approximately 870,000 emergency department visits, 1,000,000 outpatient visits, and 87,000 hospital discharges. Since the beginning of the COVID-19 pandemic, HCWs with COVID-19 symptoms had access to free-of-charge SARS-CoV-2 reverse transcriptase polymerase chain reaction (RT-PCR) testing conducted by the institution’s laboratory. During the study period, the Oxford-AstraZeneca [ChAdOx1], CoronaVac, Pfizer/BioNTech, and Janssen vaccines were available at our hospital (Supplementary Appendix 1). HCWs with a laboratory-confirmed COVID-19 infection by reverse transcriptase polymerase chain reaction (RT-PCR) were evaluated by the Employee Health Clinic (EHC) during 180 days of follow-up.

We included HCWs with a laboratory-confirmed COVID-19 infection by RT-PCR. RT-PCR testing for the diagnosis of COVID-19 was only performed on symptomatic HCWs. Cases were classified as those developing long COVID, defined as signs and symptoms that developed during or following a SARS-CoV-2 RT-PCR confirmed infection, continued for more than 4 weeks, and could not explained by an alternative diagnosis [1]. The symptoms considered related to long COVID were general symptoms (e.g., fever, tiredness or fatigue), respiratory and heart symptoms (e.g., shortness of breath, cough, chest pain, heart palpitation), neurological symptoms (e.g., headache, difficulty concentrating, change in smell or taste, depression or anxiety), digestive symptoms (e.g., diarrhea, stomach pain), or other symptoms (e.g., joint or muscle pain) [1]. HCWs that recovered from acute COVID-19 infection within four weeks were classified as controls. (Supplementary Appendix 2 and 3).

### Real-time polymerase chain reaction (RT-PCR) methodologies for SARS-CoV-2 detection

Diagnostic confirmation for COVID-19 was performed using RT-PCR on specimens obtained via nasopharyngeal swab, according to the protocol instituted at HIAE. The following RT-PCR kits were utilized: XGEN MASTER COVID-19 (Mobius, Pinhais, Paraná, Brazil), cobas®□ □SARS-CoV-2 Test (Roche Molecular Systems, Branchburg, NJ, USA), Xpert® □Xpress SARS-CoV-2 (Cepheid, Sunnyvale, CA, USA), and Abbott RealTime SARS-CoV-2 (Abbott Molecular Inc., Des Plaines, IL, USA).

### Next-generation sequencing of the viral full-length genome

We extracted total nucleic acid from naso-oropharyngeal (NOP) swab samples with the QIAamp Viral RNA Mini kit (QIAGEN, Hilden, Germany). After purification and concentration, DNAse I treatment, and depletion of human ribosomal RNA, samples were submitted to random amplification [20]. Preparation of sequencing libraries for the Illumina platform was carried out with DNA Prep (Illumina, San Diego, CA, USA) using the random two-step PCR amplification product as input. Libraries were quantified with the Qubit instrument (Thermo Fisher Scientific, Waltham, MA, USA) and loaded on the NextSeq 550 equipment (Illumina) for sequencing with MID 300 paired-end reads (Illumina).

### Exposures of interest and statistical analyses

Exposure variables were compared between those with and without long COVID-19 to identify factors associated with development of long COVID-19. Vaccination status, SARS-CoV-2 RT-PCR results of all study participants, presence or absence of long COVID, as well HCW characteristics were obtained from institutional electronic records. Baseline exposure variables included: sex, age, body mass index (BMI), physical activity (more or less than 30 minutes per day), self-reported hypertension, diabetes mellitus, arthritis, stroke, chronic kidney disease or cancer, job type (no direct patient contact or direct patient facing), the number of COVID-19 vaccine doses received prior to infection, homologous or heterologous (a booster dose different from the primary vaccine doses) COVID-19 vaccine scheme, the number of COVID-19 infections, and SARS-CoV-2 variant. HCWs were considered unvaccinated if no COVID-19 vaccine doses were received. Regarding SARS-CoV-2 variant, as only a small number of positive samples among our HCWs were sequenced, all individuals were classified in accordance to the most prevalent variant (≥ 75% of sequenced viruses) registered by the Global Initiative on Sharing Avian Influenza Data (GISAID) weekly in Brazil, at the time of the sample collection [21]. Therefore, the time period between March 2, 2020 and February 11, 2021 was considered “non-variant of concern 2020 (Non-VoC 2020) era”; February 26, 2021 to August 5, 2021 was considered the “Gamma era”; August 13, 2021 to December 16, 2021 was considered the “Delta era”; and December 24, 2021 to July 15, 2022, was considered the “Omicron era” (Figure 1).

**Figure 1.**
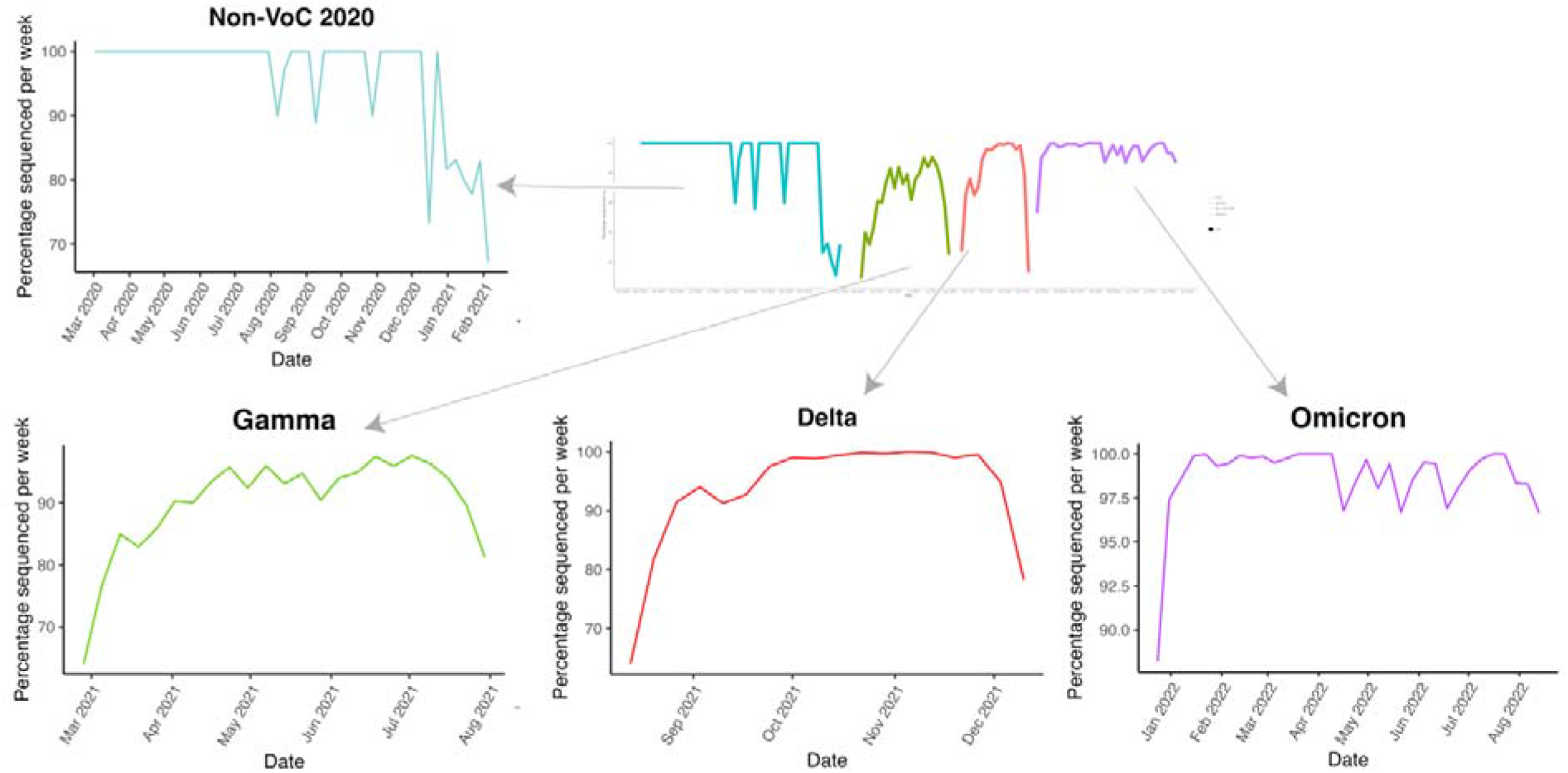
SARS-CoV-2 circulating variants.

The cumulative incidence of long COVID and its 95% confidence interval (CI) was estimated for the cohort. The univariate association between exposure variables and long COVID was assessed by Chi-Square or Fisher’s exact tests, as appropriate. Multiple logistic regression models were developed to assess the independent association between exposure variables and long COVID. Odds ratios (OR) and their respective 95% CI were calculated. Exposure variables statistically significant in the simple models were included in the multiple logistic regression models. The signs and symptoms were counted, and the relative frequencies were used to estimate correlation and build the correlation matrix. R software (v. 4.2.1) with R Studio (v. 2022.07.1) was used for the analysis. All reported tests were 2-sided and p-values <0.05 were considered significant. The study was approved by the Hospital Israelita Albert Einstein Ethics Committee (CAAE 47110421.7.0000.0071) and the need for informed consent was waived.

## RESULTS

Among a total of 18,340 HCWs at our institution, 7,051 (38.4%) HCWs had at least one laboratory-confirmed COVID-19 infection during the study period. Of those infected,1,933 (27.3%) HCWs who developed long COVID were compared to the 5,118 (72.6%) who did not (i.e., controls, Supplementary Appendix). The majority of those with long COVID had 3 or more signs or symptoms (51.8%), while 644 had one (33.3%) and 288 (14.9%) had two (Figure 2). The most common symptoms were headache (53.4%), followed by myalgia or arthralgia (46.6%), and nasal congestion (45.1%) (Figure 3, and Supplementary Appendix 2).

**Figure 2.**
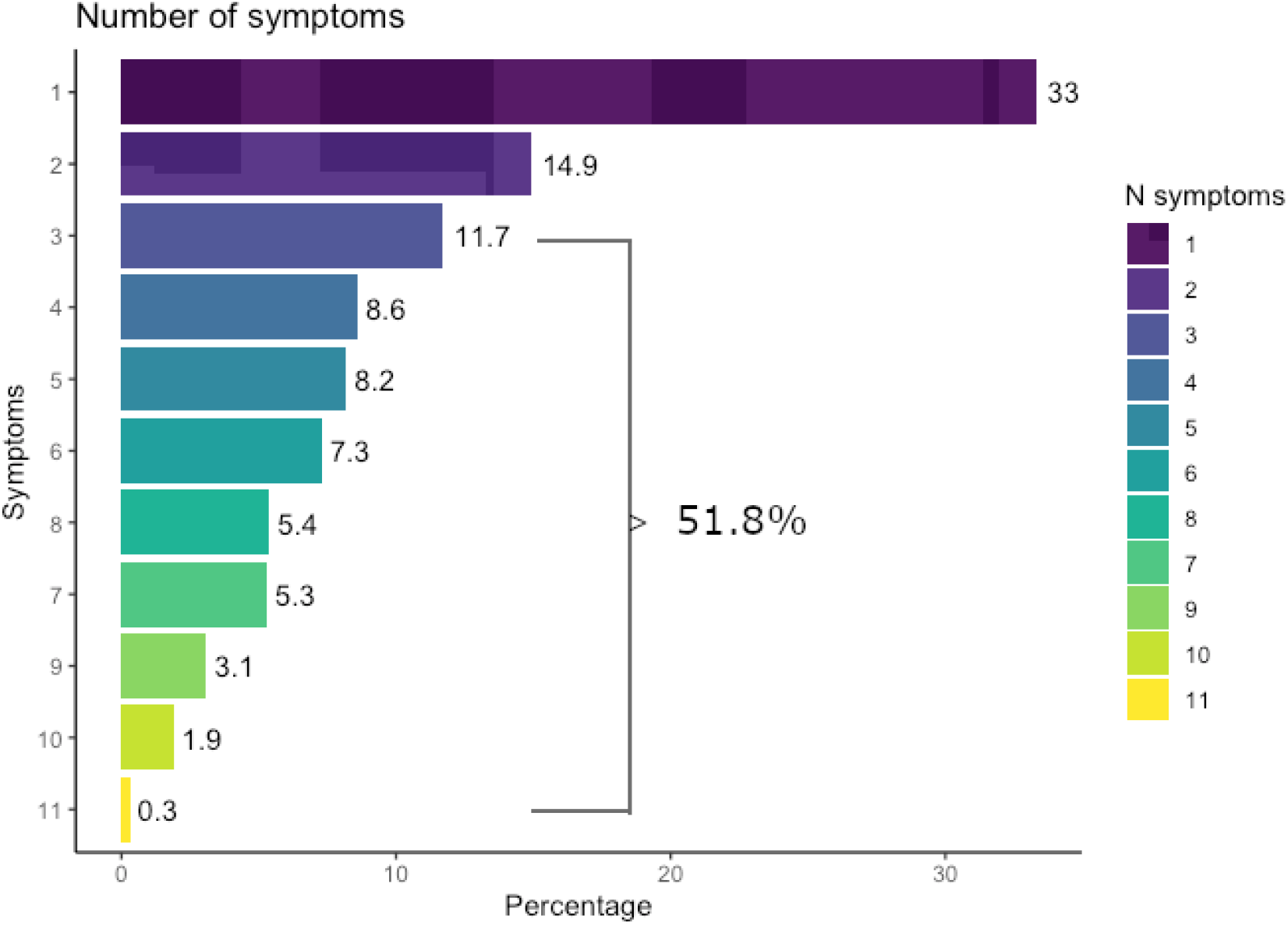
Number of long COVID signs and symptoms per HCW.

**Figure 3.**
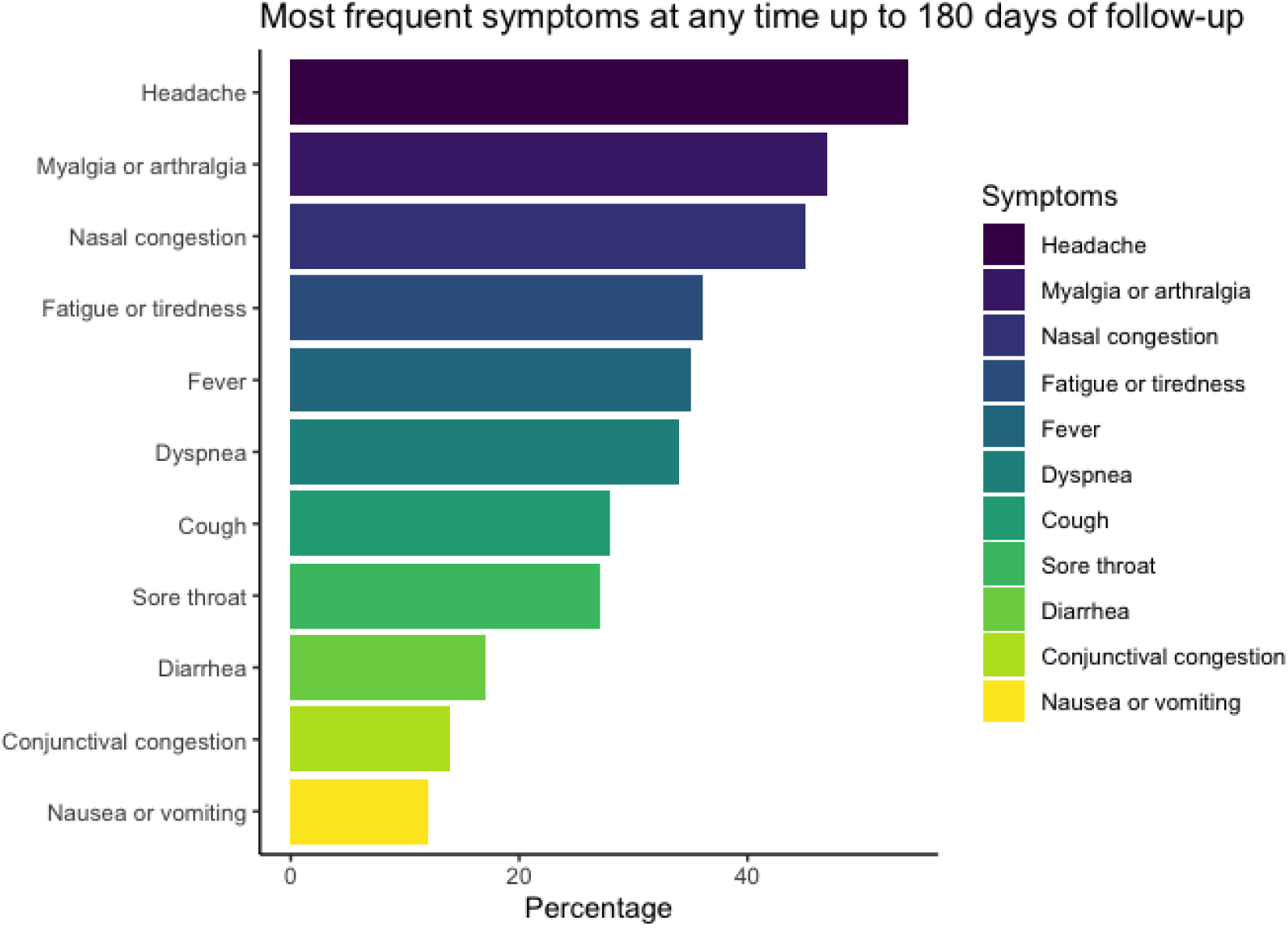
Most frequent symptoms of long COVID during 180 days of follow-up.

### Univariate analysis

Of 7,051 HCWs who had at least one COVID-19 infection, 5,101 were female (72.3%). Females represented 74.1% of the cases and 71.7% of the controls (p=0.048). The mean age for the entire infected cohort was 37.5 years, 38.1 years for those with long COVID, and 37.2 years for those without long COVID (p<0.001). Mean BMI was 27.0 overall, 27.4 for those with long COVID, and 26.8 for those without long COVID (p<0.001). Information regarding comorbidities was available for 5,722 HCWs (81.1%) in the infected cohort; at least one comorbidity was present in 28.9% overall, 31.1% in cases and 28.0% in controls (p=0.021). The most prevalent comorbidities were hypertension (9.7% in cases and 9.0% in controls), and diabetes mellitus (3.5% in cases and 2.7% in controls) (Table 1). No difference was observed in physical activity between those with and without long COVID (p=0.472), and also no difference was detected by the HCW job type (direct patient-facing or not, p=0.8). Of the infected HCWs, 3,853 (54.6%) were vaccinated before COVID-19 infection; 39.2% of cases were vaccinated (at least one dose) versus 60.5% of controls. A difference was observed in the number of vaccine doses before infection (0 to 4 doses) between those who developed long COVID and those who did not (p<0.001). The great majority of vaccinated HCWs (96.4%) received a heterologous COVID vaccine scheme and no difference was observed between cases and controls (p=0.09). Overall, 887 (12.6%) HCWs had two or more infections, including 17.8% of cases and 10.6% of controls (p=<0.001). Cases were less likely to be infected in the Omicron era (24.7%) than controls (49.8%, P<0.001).

**Table 1.** Predictors of long COVID. Variables were included in multivariable analysis if significant univariate associations were shown. The 95% confidence intervals (CIs) of the odds ratios (ORs) have been adjusted for multiple testing. Independent predictors are shown in bold.

|  | No long COVID<br>(N=5,118) | Long COVID<br>(N=1,993) | Univariate analysis |  |  | Multivariate analysis |  |  |
| --- | --- | --- | --- | --- | --- | --- | --- | --- |
|  |  |  | OR | CI 95% (II–UI) | P | OR | CI 95% (II–UI) | P |
| Female sex | 3,669 (71.7) | 1,432 (74.1) | 1.1 | (1.00-1.27) | **p=0.05** | **1.2** | **(1.05-1.39)** | **p=0.008** |
| Age, y<br>(Mean, SD) | 37.2 (9.0) | 38.1 (8.7) | 1.0 | (1.01-1.02) | **p<0.001** | **1.0** | **(1.00-1.02)** | **p=0.003** |
| BMI | 26.8 (4.8) | 27.4 (4.9) | 1.02 | (1.01-1.03) | **p<0.001** | 1.01 | (1.00-1.03) | p=0.096 |
| Any comorbidity (N = 5,722)* present | 1143 (28.0) | 512 (31.1) | 1.16 | (1.02-1.31) | **p=0.020** | 1.05 | (0.90-1.21) | p=0.5 |
| Hypertension | 365 (9.0) | 159 (9.7) | 1.09 | (0.89-1.32) | p=0.397 |  |  |  |
| Diabetes | 109 (2.7) | 58 (3.5) | 1.33 | (0.96-1.83) | p=0.084 |  |  |  |
| Arthritis | 33 (0.8) | 27 (1.6) | 2.04 | (1.22-3.41) | **p=0.006** |  |  |  |
| Stroke | 12 (0.3) | 1 (0.1) | 0.21 | (0.01-1.05) | p=0.129 |  |  |  |
| Chronic kidney failure | 8 (0.2) | 3 (0.2) | 0.93 | (0.20-3.22) | p=0.914 |  |  |  |
| Cancer | 99 (2.4) | 48 (2.9) | 1.21 | (0.85-1.70) | p=0.290 |  |  |  |
| Physical activity (N = 5,722)* -- Every days/week |  |  |  |  |  |  |  |  |
| • Never | 1454 (35.7) | 554 (33.7) |  |  |  |  |  |  |
| • 1 or 2 days | 1261 (30.9) | 520 (31.6) | 1.08 | (0.94-1.25) | p=0.273 |  |  |  |
| • 3 or 4 days | 851 (20.9) | 341 (20.7) | 1.05 | (0.90-1.23) | p=0.535 |  |  |  |
| • 5 or 6 days | 274 (6.7) | 120 (7.3) | 1.15 | (0.91-1.45) | p=0.247 |  |  |  |
| • 7 days | 237 (5.8) | 110 (6.7) | 1.22 | (0.95-1.55) | p=0.116 |  |  |  |
| Direct patient contact type job | 2637 (51.5) | 988 (51.1) | 0.98 | (0.89-1.09) | p=0.758 |  |  |  |
| Vaccine doses prior to infection |  |  |  |  |  |  |  |  |
| 0 | 2023 (39.5) | 1175 (60.8) |  |  |  |  |  |  |
| 1 | 184 (3.6) | 96 (5.0) | 0.90 | (0.69-1.16) | p=0.413 | 0.91 | (0.60-1.39) | p=0.677 |
| 2 | 665 (13.0) | 272 (14.1) | 0.70 | (0.60-0.82) | **p<0.001** | 1.17 | (0.79-1.76) | p=0.435 |
| 3 | 2115 (41.3) | 388 (20.1) | 0.32 | (0.28-0.36) | **p<0.001** | 0.63 | (0.39-1.02) | p=0.058 |
| 4 | 131 (2.6) | 2 (0.1) | 0.03 | (0.00-0.08) | **p<0.001** | **0.05** | **(0.01-0.19)** | **p<0.001** |

|  | No long<br>COVID<br>(N=5,118) | Long<br>COVID<br>(N=1,993) | Univariate analysis |  |  | Multivariate analysis |  |  |
| --- | --- | --- | --- | --- | --- | --- | --- | --- |
|  |  |  | OR | CI 95% (II-UI) | P | OR | CI 95% (II-UI) | P |
| Heterologous vaccine<br>scheme<br>administered** (N =<br>6,398) | 4498 (96.6) | 1668 (95.7) | 0.78 | (0.59-1.03) | p=0.077 |  |  |  |
| Number of COVID-19<br>infections |  |  |  |  |  |  |  |  |
| 1 | 4576 (89.4) | 1588 (82.2) |  |  |  |  |  |  |
| ≥2 | 542 (10.6) | 345 (17.8) | 1.83 | (1.58-2.12) | **p<0.001** | **1.27** | **(1.07-1.50)** | **p=0.006** |
| Variant Era***<br>(N=6,918) |  |  |  |  |  |  |  |  |
| Non-VoC 2020 | 1,930 (38.4) | 1,133 (59.8) |  |  |  |  |  |  |
| Delta | 214 (4.3) | 43 (2.3) | 0.34 | (0.24-0.47) | **p<0.001** | **0.30** | **(0.17-0.50)** | **p<0.001** |
| Gamma | 377 (7.5) | 250 (13.2) | 1.13 | (0.95-1.35) | p=0.174 | 1.03 | (0.72-1.48) | p=0.9 |
| Omicron | 2502 (49.8) | 469 (24.7) | 0.32 | (0.28-0.36) | **p<0.001** | **0.49** | **(0.30-0.78)** | **p=0.003** |

\*Information available for 5,722 (81.1%)
\*\*Any heterologous COVID-19 vaccine scheme
SD=Standard deviation
BMI=Body mass index
\*\*\*113 HCWs were not classified due to potential overlap between Non-VoC and Gamma, Gamma and Delta, and Delta and Omicron variants.

### Multivariable analysis

In the multivariable analysis assessing the association between exposure variables and long COVID, female sex (OR 1.21 [CI95v 1.05-1.39]), older age (OR 1.01 per year of age [CI95 1.00-1.02]), and two or more COVID-19 infections (OR 1.27 [CI95 1.07-1.50]) were significantly associated with development of long COVID. Those infected with the Delta variant (OR 0.30 [CI95 0.17-0.50]) or the Omicron variant (OR 0.49 [CI95 0.30-0.78]), and those who received 4 vaccine doses before infection (OR 0.05 [CI95 0.01-0.19]) were associated with a reduced risk of development of long COVID (Table 1 and Supplementary Appendix 4).

### Whole-genome sequencing analysis

During the study period, SARS-CoV-2 samples from 524 (6.6%) of the 7,979 RT-PCR positive test results were sequenced to determine the virus variant. Most of the samples were Delta (45.6%), followed by Omicron (31.0%), and Gamma (20.8%). Alpha, B.1, B.1.1, B.1.1.28 and Zeta accounted for less than 1%. And 4 samples were inconclusive (0.8%) (Supplementary Appendix 5). The agreement between results of whole-genome sequencing analysis and the circulating variant eras that we defined was 438/518 (84.6%).

## DISCUSSION

In this case-control study of HCWs from Brazil, more than one quarter (27%) had long covid symptoms in the 6 months following infection. Acquiring more than one COVID-19 infection was associated with a higher risk of long COVID. The most common long COVID symptom was headache and about a half of those with long COVID-19 had more than 2 symptoms. Risk factors associated with development of long COVID-19 were female sex, age, and two or more COVID-19 infections. Protective factors were the Delta variant, the Omicron variant, and receiving four doses of COVID-19 vaccine prior to infection.

A recent systematic review and meta-analysis demonstrated that long COVID is a public health issue with an overall global estimated pooled prevalence of 43% (95% CI, 39%-46%) with a prevalence of 54% (95% CI, 44%-63%) in hospitalized patients and 34% (95% CI, 25%-46%) in non-hospitalized patients [2]. Another systematic review including 57 studies reported that more than half of COVID-19 survivors experienced persistent post-COVID condition symptoms 6 months after recovery [22]. Multiple papers have reported a variety of symptoms and durations to make a diagnosis of long COVID [2, 22-25]. The most common symptoms described in previous papers were fatigue or muscle weakness, persistent muscle pain, anxiety, memory problems, sleep problems, and shortness of breath [2, 22, 24]. In our case-control study with up to 180 days of follow-up, headache, myalgia/arthralgia, nasal congestion, and fatigue were the most common symptoms. A previous paper from UK reported the major risk factors for not feeling fully recovered at 1 year after COVID-19 were female sex, obesity, and receiving invasive mechanical ventilation during the acute illness [25]. Another study reported that regardless of the initial disease severity, COVID-19 survivors had longitudinal improvements in physical and mental health, with most returning to their original work within two years [24]. In the multivariate analysis of our study, females were also more affected by long COVID than males. The reason for this is not clear but has also been shown in previous studies [23, 26, 27].

Our study revealed that more than one COVID-19 infection was associated with long COVID-19 among HCWs. Previous evidence has shown that COVID-19 reinfection can increase the risk of having long-term health complications including all-cause mortality, hospitalization, and post-acute sequelae [10]. More attention should be given to COVID-19 infection among HCWs particularly when there’s a previous history of infection (i.e., reinfection) and vaccination status [10].

The SARS-CoV-2 VoC have multiple spike protein mutations and appear to be more infectious or cause more disease than other circulating variants [28]. A growing body of early global research shows a clear effect of the vaccines against the new variants [23, 29-31]. Previous studies suggested that the Delta and Omicron variants caused less systemic inflammatory processes, severe illness, or death, resulting in less severe long COVID symptoms than the wild-type variant (Wuhan) [32, 33]. In addition, the prevalence of long COVID during the Omicron era has been reported as less than that of the other strains, and a milder process during the acute phase might have been contributing to the less frequent development of long COVID during the Delta and Omicron eras [33, 34].

The protective effect of COVID-19 vaccination versus long COVID is not well described. A recent systematic review and meta-analysis demonstrated that COVID-19 vaccination of at least one dose either before or after having COVID-19 infection significantly decreased the risk of long COVID [35]. Recently, Azzolini et al. demonstrated receiving 2 or more doses of COVID-19 vaccines before COVID-19 infection was associated with a lower prevalence of long COVID [16]. However, this present study revealed that receiving 4 doses of COVID-19 vaccinations before infection was a protective against long COVID, although a dose-response effect was not detected, which could have been due to the small sample size, creating an unbalanced comparison between vaccinated and unvaccinated. Vaccinations may play a role in preventing long COVID and further studies need to be conducted with newer COVID-19 vaccinations included.

Our study had several limitations. First, we did not perform a test-negative case-control study design because this study was retrospectively conducted using the data from symptom-based testing and all included subjects had documented COVID infection. Second, there is a possibility that HCWs had COVID-19, developed long COVID, but did not come back to employee health, therefore were considered as not having long COVID leading to misclassification of the outcome. Third, since our institution offered COVID-19 testing for only symptomatic HCWs, we did not include those with asymptomatic COVID-19, although long COVID can develop following asymptomatic infection [1]. Fourth, a clearer and more standardized definition of long COVID is needed for researchers to investigate the true prevalence and to better evaluate the risk factors since there is no test to diagnose long COVID [1, 36]. Fifth, neutralizing viral antigen-binding antibody levels were not available in the vaccinated HCWs in our study. However, the U.S. FDA does not recommend antibody testing for SARS-CoV-2 to determine immunity or protection from COVID-19, especially among those who are vaccinated [37]. Sixth, past medical history was not available for approximately 20% of the study subjects. We also could not perform further analyses stratified by immunocompromised status due to the limited number of individuals (<3%) reported in our previous study [38]. Since our study focused only on long COVID among HCWs, we were not able to evaluate the impact of personal protective equipment. Finally, the most prevalent variant at the time of infection was used to assess long COVID risk since viral sequencing was limited to a small random sample of 524 HCWs, corresponding to 7% of HCWs diagnosed with COVID-19 included in this study.

However, the agreement between results of whole-genome sequencing analysis and the circulating variant eras that we defined wwas about 85%.

## Conclusions

Our study demonstrated that long COVID can be prevalent among HCWs. Importantly, multiple infections were associated with increased risk of long COVID, and the more recent Delta and Omicron variants were associated with reduced risk. COVID-19 vaccines were significantly associated with reduced long COVID among HCWs after additional doses (second booster). More studies are needed to evaluate long COVID after reinfection and additional genomic surveillance is required for a better understanding of vaccine effectiveness vs long COVID after infection with newer SARS-CoV-2 variants.

## Data Availability

All data produced in the present work are contained in the manuscript

## Acknowledgements

We thank all the participants for their contributions to this study.

## Data sharing agreement

De-identified individual study data reported in this article can be requested upon application to investigator board via the corresponding author.

