## Supplementary Appendix for "Risk factors for long COVID among healthcare workers, Brazil, 2020–2022"

**Supplementary Appendix 1.** COVID-19 vaccine schemes among 7,051 healthcare workers (HCWs)*

|  |  | **COVID-19 vaccine first dose N (%)** | | |  |
| --- | --- | --- | --- | --- | --- |
|  |  | CoronaVac | [ChAdOx1] | Janssen | Pfizer/BioNTech |
| **COVID-19 vaccine second dose N (%)** | CoronaVac | 3,256 (100) | - | - | - |
|  | [ChAdOx1] | - | 3,209 (98.7) | - | - |
|  | Pfizer/BioNTech | - | 41 (1.3) | - | 148 (100) |
|  | Janssen | - | - | 5 (100) | - |

Oxford-AstraZeneca [ChAdOx1]

*392 unvaccinated HCWs (5.6%), 6,659 vaccinated HCWs (94.4%)

5,124 had one booster and 1,284 had two boosters COVID-19 vaccines

**Supplementary Appendix 2.** Associated symptoms to the Long COVID

| **Symptoms** | **N (%)** |
| --- | --- |
| Headache - Yes | 1,123 (53.4) |
| Myalgia or arthralgia | 980 (46.6) |
| Nasal congestion | 948 (45.1) |
| Fatigue or tiredness | 744 (35.4) |
| Fever | 736 (35.0) |
| Dyspnea | 716 (34.0) |
| Cough | 602 (28.6) |
| Sore throat | 592 (28.1) |
| Diarrhea | 345 (16.4) |
| Conjunctival congestion | 293 (13.9) |
| Nausea or vomiting | 238 (11.3) |

**Supplementary Appendix 3.** Flowchart

No Long COVID (n = 5,118)

Long COVID (n = 1,933)

Laboratory confirmed COVID-19 infection

(n = 7,051)

Assessed for eligibility

(n = 18,340)

Excluded (n= 11,289)

♦  No laboratory confirmed COVID-19 test or symptoms

**Supplementary Appendix 4.** Multivariate analysis of predictors of long COVID.

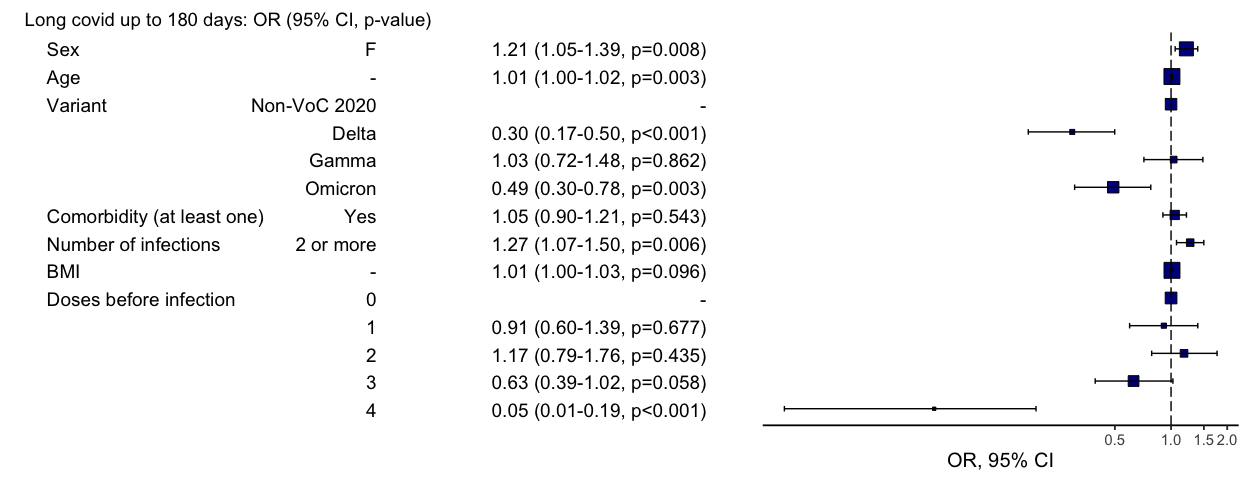

**Supplementary Appendix 5.** SARS-CoV-2 virus lineages sequenced (n=524)

| **Lineage** | **N** | **%** | **Agreement with dominant strain* (%)** |
| --- | --- | --- | --- |
| Alpha | 1 | 0.2 | 0/1 (0) |
| B.1 | 1 | 0.2 | 1/1 (100) |
| B.1.1 | 2 | 0.4 | 1/2 (50) |
| B.1.1.28 | 3 | 0.6 | 1/3 (33) |
| Delta | 239 | 45.6 | 213/239 (89.1) |
| Gamma | 109 | 20.8 | 88/109 (80.7) |
| Omicron | 163 | 31 | 134/163 (82.2) |
| Zeta | 2 | 0.4 | 0/2 (0) |
| Inconclusive | 4 | 0.8 |  |
| **Total** | 524 | 100.0 |  |

*% of agreement between the SARS-CoV-2 virus genomes sequenced with the dominant variant at the time the sample was obtained: 438/518 (84.6%).

.
